# Forecasting Trajectories of an Emerging Epidemic with Mathematical Modeling in an Online Dashboard: the Case of COVID-19

**DOI:** 10.1101/2020.05.21.20108753

**Authors:** Wojciech Młocek, Robert Lew

**Affiliations:** University of Agriculture in Krakow, Balicka 253c, 30-198 Kraków, Poland; Adam Mickiewicz University in Poznań, Grunwaldzka 6, 60-780 Poznań, Poland

## Abstract

We offer an efficient mathematical model for forecasting the course of an emerging epidemic, with COVID-19 as a use case. We predict the future course of confirmed cases in a number of countries, and present the results in a modern online dashboard, updated daily and accessible to the public.

## 1 Introduction

The 2019/2020 COVID-19 pandemic, caused by the SARS-CoV-2 coronavirus, and believed to have had its source in Wuhan, China [1, 2], has come to be seen as a major public health emergency in numerous countries. Its dynamic spread has prompted authorities to introduce a broad variety of measures, and has mobilized resources on a heretofore unprecedented scale, as well as attracted wide attention among experts, politicians, and the general public [3]. In planning the measures and resources, it is crucial that timely and reliable data are available on the current prevalence of the disease, and – more pertinently to the present contribution – on its future course [4, 5]. Forecasts of the future course of the disease, as expressed through the trajectory of cases (and, ideally, their severity), are essential in the planning of the extent and nature of epidemic response. Further, given the enormous and widespread interest in the topic, such data should be amenable to online presentation, in a form that is both visually attractive and easy to assimilate by non-epidemiologists.

An emerging epidemic presents particular challenges to attempts at predicting its future course for a number of reasons. For lack of space, let us just list the following:

- the characteristics of a new virus are largely unknown;
- a wide range of non-pharmacological interventions are introduced (and withdrawn), whose impact on epidemic spread is not known (containment strategies like contact tracing, isolation and crowd limitations);
- initial reporting may be incomplete, delayed, or inconsistent (for example, what exactly is reported: individuals, tests sampled, tests performed, specimens, individual swabs? At the time of this writing, it is still often unclear which is the case [6]);
- testing policy is subject to change with respect to both testing intensity and selection of tested individuals;
- the sensitivity and specificity of early tests is not well known as there is no reliable gold standard; and, tests may vary markedly on these parameters.

Given the extent of the COVID-19 crisis, its volatility, uncertainty, and amount of confusion caused, availability of up-to-date, credible epidemic projections is of the essence, ideally available in an accessible online format.

To respond to this urgent need, with this contribution we propose and implement a mathematical model for forecasting the course of an emerging epidemic, with COVID-19 as our use case. We apply this model to predict the future course of confirmed cases in a number of countries, and present the results in a modern, interactive online dashboard, updated daily and accessible to the public. The approach includes a robust, real-time validation procedure which consists in replicating past projections and comparing them with actual observed data (see Subsection 2.3). Unlike in most compartmental (SIR, SEIR, and further extensions) approaches [7–9], our models do not require prior estimation of the usual epidemic pathogen-specific parameters (R_0_, incubation period), which presents a particular challenge for emerging pathogens and diseases, such as the SARS-CoV-2 and COVID-19 [5, 10–15]. Further, our approach can be easily applied to levels other than country (e.g. state, county, province), and to data other than confirmed cases (e.g. deaths, hospitalizations, or severe cases). Details of the approach are set out in Section 2. In Section 3, we shall proceed to explain actual forecasts for selected countries.

## 2 Method

### 2.1 Modeling the case trajectories

Let *z*: [0, +∞) → ℝ be a continuous function. Assume further that the *z* function describes the progression of the epidemic, with *z*(*t*) representing the cumulative number of confirmed infections at time point *t*. The graphic representation of this function is often referred to as the *epidemic curve*. Let’s also assume that time is a continuous variable, although in practice we will measure it in daily increments, representing the day-by-day course of the epidemic. If *z*(*t*) represents the total number of confirmed infections (i.e. ’cases’) on day t, then *z′*(*t*) will be the rate of change of the epidemic, being the number of new cases on day *t*. Assume that the ratio of new cases on a given day to total cases is described by the function *u*, thus:

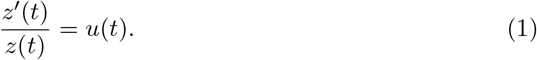

We obtain, then, the following linear differential equation

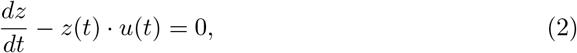

with a general solution of the form:

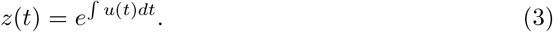

Therefore, in order to be able to predict the course of the epidemic, we need the *u* function, which could be defined in a number of ways. Here, we assume that the ratio of new cases to total cases will decrease exponentially, thus:

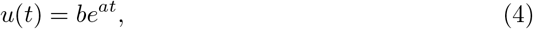

where *a <* 0, and *b* > 0. The *b* parameter expresses the ratio of new cases to total cases at day 0, while the *a* parameter describes the rate of decline of the ratio. Inserting the function *u* expressed as (4) into Eq (3), we obtain the *z* function, which describes the progression of the epidemic:

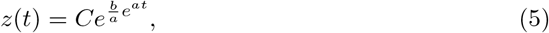

where *C* > 0. The above is one variant of the Gompertz growth equation [16]. Note that since *a* < 0, then 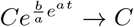 at *t* → +∞, thus *C* represents the maximum number of cases.

### 2.2 Estimation of model parameters

To estimate model parameters, we use the latest confirmed cases data as made available by the Center for Systems Science and Engineering (CSSE) at Johns Hopkins University (JHU)(https://github.com/CSSEGISandData/COVID-19).

Let the time series *z*_0_, *z*_1_,…,*z_T_* be the cumulative number of confirmed cases and 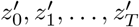 be the number of new cases at times 0, 1,…,*T*. Time *T* is the day of the forecast. Take, day zero” to be the first day on which the cumulative number of cases has reached at least q% of the total cases on the day of the forecast, that is at least *q·z*_T_. Let’s refer to the number *q* as the *cutoff level*, as it cuts off the early stage of the epidemic. We initially assume *q* = 5%. Should it turn out that, on the day of the forecast, neither model is adequate, i.e. the maximum number of cases *C* < *z_T_*, we increment the cutoff level *q* by the lowest number (to the nearest 1%) at which, for at least one model

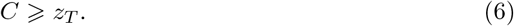

We estimate the model parameters numerically using the Levenberg-Marquardt (L-M) algorithm [17, 18], also known as the *damped least-squares* approach. We do this in three ways, referred to, for convenience as Model 1, Model 2 and Model 3, respectively.

Model 1.

We first do a log transformation

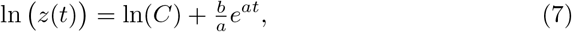

and then we apply the L-M algorithm to points ln *z*_0_, ln *z*_1,…,_ln *z_T_*.

Model 2.

Here we apply the L-M algorithm directly to points *z*_0_, *z*_1_,…,*z_T_*, getting the starting estimates from Model 1.

Model 3.

We apply the L-M algorithm to points defined as 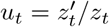, *t* = 0, 1,…,*T*, for *u* given in (4). In this way, we estimate parameters *a* and *b*. Assuming that the solution of Eq (2) given in (5) meets the initial condition of *z*(*T*) = *z_T_*, we obtain:

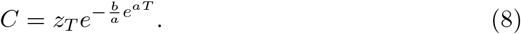

The difference between the three approaches to parameter estimation brings into focus the influence of daily variation in new cases on the model parameters. Model 1 is most dependent on the early values used in the forecast, since a logarithmic function becomes less steep with time. Model 2 is equally sensitives to values along the whole range. So is model 3, but in this case we add the condition of perfect accuracy on the day of the forecast, that is *z*(*T*) = *z_T_*. These differences will be reflected in the shapes of the epidemic curves. Condition (6) will only be checked for Models 1 and 2, since it is met by definition for Model 3.

To evaluate the fit of the model (i.e., quality of the forecast), we compute the following common statistical measures of fit for observed *z_t_* and predicted *z*(*t*) cases:

- The Nash–Sutcliffe model efficiency coefficient [19]

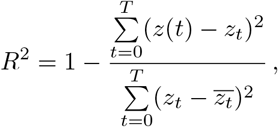

where 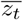 is the mean of observed cases. It is also known as the coefficient of determination.
- Percentage error (PE)

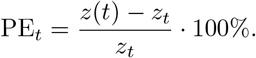
- Mean absolute percentage error (MAPE)

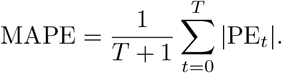

### 2.3 Model validation

To validate the model, we go back ten days from the day of the forecast. Thus, if the forecast is made on the 50th day of the epidemic, we go back to day 40, and, at day 40, we compute predictions for 7 consecutive days (these being days: 41, 42, 43, 44, 45, 46, and 47 of the epidemic). For each of these, we compute the percentage error (PE). Then we move forward by one day to day 41 of the epidemic and repeat the above steps, computing the predictions, at day 41, for the 7 consecutive days (which at this point are epidemic days: 42, 43, 44, 45, 46, 47, and 48), and again compute the PE for each of the seven days. We continue advancing by one day in this way. Note that, starting with day 44, the forecasts will no longer cover a complete week (as we only generate the predictions for the days on which we have actual observations, that is up to day 50). In this case, the final prediction will be on day 49 of the epidemic, where we only predict the value for day 50. The PE obtained in this way are presented as box plots, and they yield the mean absolute percentage error (MAPE). This serves as a measure of the current usefulness of the model.

## 3 Results and Discussion

Our forecasts are computed daily from data made available by the Center for Systems Science and Engineering at Johns Hopkins University (https://github.com/CSSEGISandData/COVID-19), and published daily on an interactive dashboard (see Section 4). Here, for ease of exposition, we present COVID-19 epidemic forecasts as computed specifically on **May 14, 2020**. We computed predictions for the following countries: the US, Italy, Germany, and the United Kingdom (Fig 1); and Mexico, Poland, Sweden, Turkey (Fig 2). Figs 1-2 present the predicted numbers of confirmed COVID-19 cases (up to May 14, 2020) and predicted epidemic curves for a prediction horizon of 100 days.

**Fig 1.**
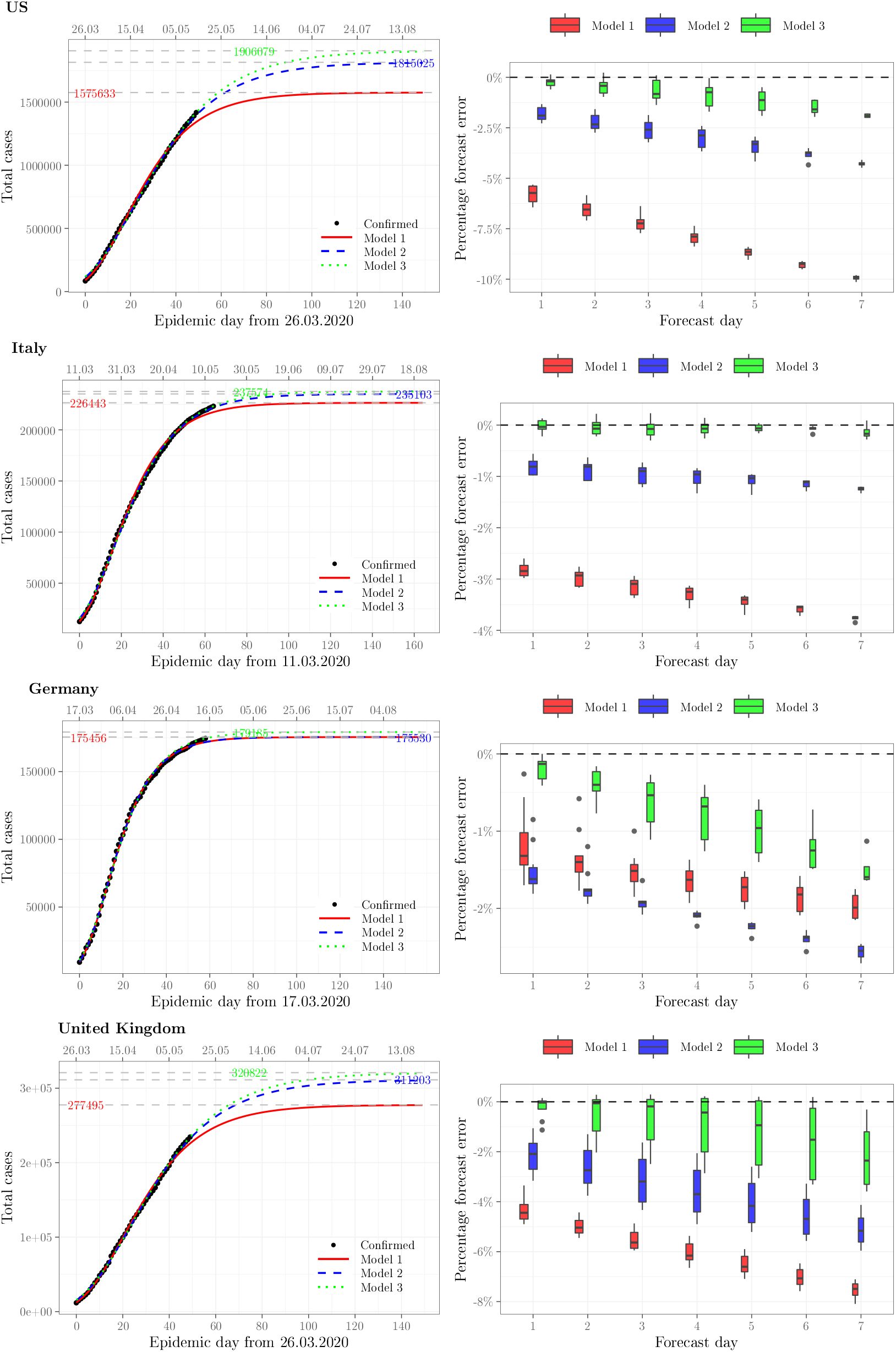
Observed and projected COVID-19 epidemic curves (left) and corresponding box plots of percentage forecast error (right) for the US, Italy, Germany and the United Kingdom

**Fig 2.**
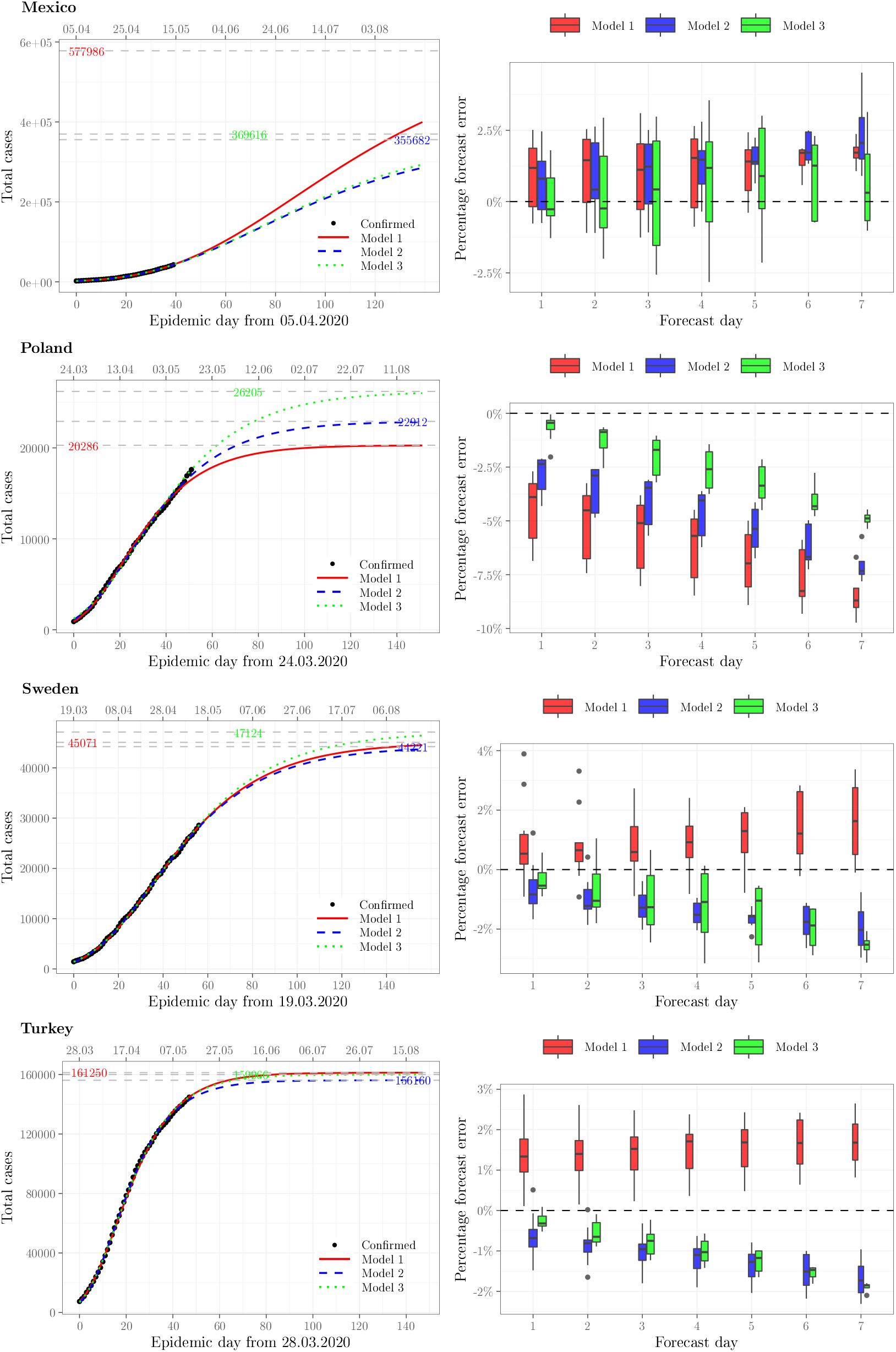
Observed and projected COVID-19 epidemic curves (left) and corresponding box plots of percentage forecast error (right) for Mexico, Poland, Sweden and Turkey

Measures of fit and prediction accuracy display a very close match between the predicted values and observed cumulative confirmed cases (see Table 1). The coefficient of determination (*R*^2^) is close to 100% for all countries, while the mean absolute percentage error (MAPE) for most countries is less than 3 percent. The largest prediction error was found for Poland in Model 3 (MAPE = 6.19%).

**Table 1.** *R*_2_ as a measure of model fit and MAPE as a measure of prediction accuracy (selected countries)

| Country | Model 1 |  | Model 2 |  | Model 3 |  |
| --- | --- | --- | --- | --- | --- | --- |
| | $R^2$ [%] | MAPE [%] | $R^2$ [%] | MAPE [%] | $R^2$ [%] | MAPE [%] |
| Germany | 99.91 | 1.62 | 99.91 | 1.64 | 99.86 | 2.46 |
| Italy | 99.78 | 1.96 | 99.90 | 3.03 | 99.90 | 3.26 |
| Mexico | 99.94 | 1.83 | 99.96 | 2.18 | 99.96 | 2.11 |
| Poland | 99.67 | 2.18 | 99.82 | 4.14 | 99.67 | 6.19 |
| Sweden | 99.95 | 2.00 | 99.95 | 2.02 | 99.93 | 2.33 |
| Turkey | 99.92 | 1.58 | 99.96 | 1.78 | 99.94 | 1.61 |
| United Kingdom | 99.75 | 2.05 | 99.89 | 3.56 | 99.88 | 3.90 |
| US | 99.59 | 2.78 | 99.88 | 3.89 | 99.85 | 4.49 |

To answer the question as to which of the three scenarios is more likely for a given country, it is helpful to examine the percentage errors of the three forecasts. Figs 1-2 give the statistical characteristics of forecast errors for the three Models. Percentage errors are computed for forecasts starting ten days back, that is starting on May 4, 2020. Our box plots (Figs 1–2) indicate the range of the errors, as well as the under- or overestimation of the cumulative confirmed cases. For instance, we see that for the US, Germany, United Kingdom, and Poland, all three models tend towards underestimation, which would suggest a pessimistic scenario. By contrast, we see some overestimation for Mexico, which suggests a more optimistic scenario given by Models 2 and 3. Yet another group of cases are Sweden and Turkey, where Model 1 overestimates, and Models 2 and 3 underestimate; here, it is hard to pick the more likely scenario.

In addition to the box plot visualization of the percentage of forecast error, Table 2 gives the mean absolute percentage forecast errors (MAPE) for each of the forecast days 1, 2,…,7, separately and jointly (1–7). The heat map version of the table lets the reader see at a glance which country projections are the most and least accurate. In most cases, it is Model 3 that yields best weekly forecasts. This follows from the fact that Model 3 has, by definition, 100% accuracy on the day of the forecast. We should stress that the three models presented here are characterized by the same Eq (5), and they reflect well the evolution of the epidemic in time by emphasizing the different effect of epidemic stages on their parameters. Trajectory changes reflect changes to the epidemic dynamic in a given country. For instance, for the US, Italy, the United Kingdom, and especially for Poland, we note a transition to a more pessimistic trajectory (Model 3) compared to that followed in the early stages (Model 1). By contrast, Mexico displays the reverse tendency, which could be taken to suggest a reduction in the dynamic of the epidemic in that country.

**Table 2.** Mean absolute percentage error (MAPE [%]) for forecast days 1 to 7 (selected countries)

| Country | Forecast days (Model 1) |  |  |  |  |  |  |  | Forecast days (Model 2) |  |  |  |  |  |  |  | Forecast days (Model 3) |  |  |  |  |  |  |  |
| --- | --- | --- | --- | --- | --- | --- | --- | --- | --- | --- | --- | --- | --- | --- | --- | --- | --- | --- | --- | --- | --- | --- | --- | --- |
|  | 1 | 2 | 3 | 4 | 5 | 6 | 7 | 1–7 | 1 | 2 | 3 | 4 | 5 | 6 | 7 | 1–7 | 1 | 2 | 3 | 4 | 5 | 6 | 7 | 1–7 |
| Germany | 1.2 | 1.4 | 1.5 | 1.6 | 1.8 | 1.8 | 2.0 | 1.5 | 1.5 | 1.7 | 1.9 | 2.1 | 2.2 | 2.4 | 2.6 | 2.0 | 0.2 | 0.4 | 0.6 | 0.8 | 1.0 | 1.2 | 1.5 | 0.7 |
| Italy | 2.8 | 3.0 | 3.1 | 3.3 | 3.4 | 3.6 | 3.8 | 3.2 | 0.8 | 0.9 | 1.0 | 1.0 | 1.1 | 1.1 | 1.2 | 1.0 | 0.1 | 0.1 | 0.1 | 0.1 | 0.1 | 0.1 | 0.2 | 0.1 |
| Mexico | 1.3 | 1.4 | 1.4 | 1.4 | 1.3 | 1.4 | 1.7 | 1.4 | 1.1 | 1.2 | 1.3 | 1.4 | 1.5 | 1.9 | 2.4 | 1.4 | 0.8 | 1.4 | 1.8 | 2.0 | 1.7 | 1.4 | 1.5 | 1.5 |
| Poland | 4.4 | 5.0 | 5.6 | 6.2 | 6.9 | 7.7 | 8.5 | 6.0 | 2.8 | 3.4 | 4.0 | 4.7 | 5.4 | 6.2 | 7.0 | 4.4 | 0.6 | 1.2 | 2.0 | 2.6 | 3.3 | 4.0 | 4.9 | 2.3 |
| Sweden | 1.1 | 1.1 | 1.1 | 1.2 | 1.3 | 1.5 | 1.7 | 1.2 | 0.9 | 1.1 | 1.3 | 1.5 | 1.7 | 1.8 | 1.9 | 1.4 | 0.6 | 1.0 | 1.3 | 1.3 | 1.5 | 2.0 | 2.6 | 1.3 |
| Turkey | 1.4 | 1.4 | 1.4 | 1.5 | 1.6 | 1.6 | 1.7 | 1.5 | 0.7 | 0.8 | 1.0 | 1.2 | 1.4 | 1.5 | 1.7 | 1.1 | 0.3 | 0.5 | 0.8 | 1.0 | 1.3 | 1.6 | 1.9 | 0.9 |
| United Kingdom | 4.3 | 5.0 | 5.5 | 6.0 | 6.5 | 7.0 | 7.5 | 5.7 | 2.1 | 2.6 | 3.1 | 3.6 | 4.0 | 4.5 | 5.1 | 3.3 | 0.3 | 0.6 | 0.9 | 1.1 | 1.4 | 1.7 | 2.1 | 1.0 |
| US | 5.8 | 6.5 | 7.2 | 7.9 | 8.7 | 9.3 | 9.9 | 7.5 | 1.8 | 2.2 | 2.6 | 3.0 | 3.4 | 3.8 | 4.3 | 2.8 | 0.3 | 0.5 | 0.7 | 0.9 | 1.2 | 1.5 | 1.9 | 0.8 |

## 4 The Interactive Dashboard

To be of maximum utility, our models should be calibrated daily. In order to facilitate that, we have created an online dashboard, which allows any interested parties to follow, in real time, daily forecasts for dozens of countries. The dashboard affords an interactive view of the projected epidemic trajectories, supplemented with measures of model fit and prediction error. The dashboard, available at the following URL: https://rebrand.ly/covid_dashboard is produced in the R environment [20] for statistical computing, enhanced with rmarkdown, flexdashboard and plotly packages [21–23], and uploaded to a WWW server. Upon loading, the dashboard displays the forecast (our three Models) as a line plot of predicted cumulative confirmed cases, overlaid on a dot plot of observed daily case totals, for a default country (currently: the US). A screenshot is presented in Fig 3.

**Fig 3.**
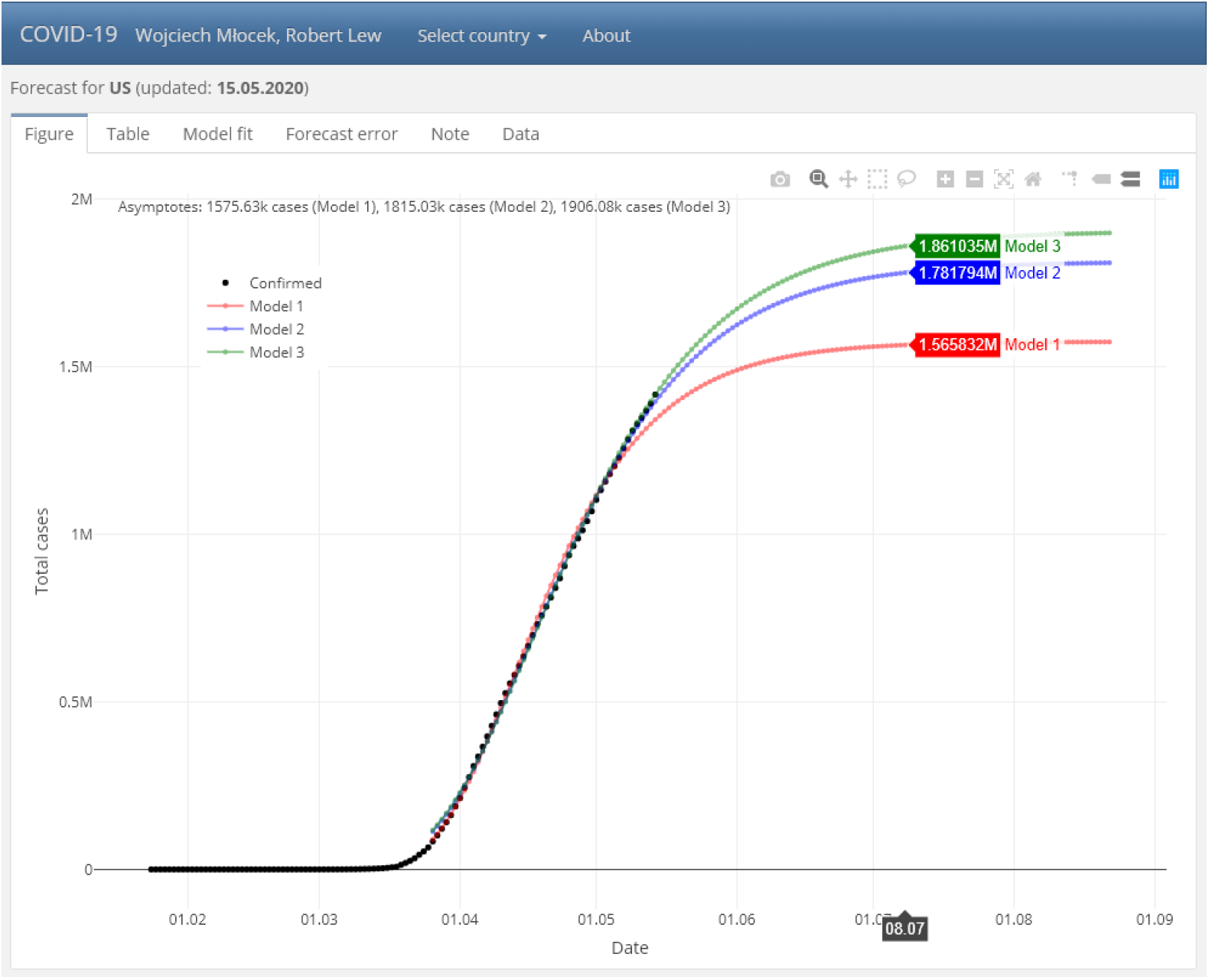
A screenshot of the interactive dashboard available at https://rebrand.ly/covid_dashboard

Further countries are selected from the top-level menu, using a drop-down control, arranged alphabetically. An *About* section references this submission to PLoS ONE (and will be replaced by the final citation, should the article be accepted for publication).

A second-level menu applies to the country selected, and consists of six tabs:

1. the *Figure* tab presents four time series line plots, overlaid: observed confirmed cases and forecasts as per the three Models;
2. the *Table* tab gives the table of daily predicted cumulative confirmed case counts;
3. the *Model fit* tab gives the graphic representation of model fit with statistical measures of fit;
4. the *Forecast error* tab has box plots of percentage forecast errors for the three models, each on seven days, as well as the mean absolute percentage error (MAPE);
5. the *Note* tab offers some documentation, including the exact equations for the three predicted curves;
6. finally, the *Data* tab tabulates the observed daily cumulative confirmed cases and daily new cases, referencing the source.

We plan to develop the dashboard further while maintaining the current functionality. We have also created a sister dashboard for individual US States, available at: https://rebrand.ly/covid_dashboard_US.

## 5 Conclusion

A crucial aspect in planning the strategy of countering the COVID-19 epidemic is forecasting the spread of the SARS-CoV-2 pathogen, both short- and long-term. The approach presented in the present contribution makes it possible to predict accurately new cases in the coming week. Beyond that, it also offers viable scenarios of epidemic spread for weeks and months to come. Thanks to the accuracy and daily calibration of the models, they follow the epidemic path closely. It is hoped that the forecasts presented in an interactive, updated dashboard will be highly useful in managing the COVID-19 pandemic effectively, and the approach can be extended to future epidemic events.

## Data Availability

We use the latest confirmed cases data made available by the Center for Systems Science and Engineering (CSSE) at Johns Hopkins University (JHU)

https://github.com/CSSEGISandData/COVID-19

